# Singular Value Decomposition-Based Coil Combination Improves the Accuracy and Noise-Robustness of Quantitative Susceptibility Maps

**DOI:** 10.64898/2026.05.28.26354148

**Authors:** Caleb Atkins, Tianxia Wu, Brandon Bujak, Souheil Inati, Peter Kellman, Govind Nair

## Abstract

Most high-field MRI scanners conduct imaging using phased-array coils, in which the signals received by an array of coil elements are combined for downstream processing. Optimally combining these signals requires knowledge of each coil’s spatial sensitivity profile, which can be acquired from a volume coil with homogeneous sensitivity across the field-of-view. However, this approach is not often used on high-field MRI scanners, especially on non-clinical systems; therefore, this work uses an algorithm based on the singular-value decomposition (SVD), called SVD-B1, to estimate coil sensitivities directly from the array data itself. Images produced by SVD-B1 are devoid of wormhole artifacts and open-ended fringe lines commonly seen in more conventional reconstructions. Quantitative Susceptibility Maps (QSMs) produced using the algorithm were compared to those produced using other combination algorithms across clinically relevant regions of in-vivo and postmortem human brains. As progressive levels of simulated noise were added to the data, SVD-B1’s QSMs were up to 3% (in-vivo) and 13% (postmortem) more consistent (as measured by their Intraclass Correlation Coefficient) than those from other algorithms. Additionally, these QSMs were up to 8.5% (in-vivo) and 36% (postmortem) more accurate than other QSMs with respect to a “single-coil” reference. A parallel imaging extension of SVD-B1, called SVD-B1 GRAPPA, achieved similar results for QSMs generated from progressively more accelerated acquisition data. These results show that SVD-B1 can improve the sensitivity of high-resolution QSM to subtle changes in fine-grained tissue structures (e.g., in neurodegenerative disease) and help reduce scan times in clinical settings where shorter scans are imperative.

## Introduction

Most radiofrequency (RF) receive coils used in high-field MRI follow a phased-array (PA) design, involving multiple, smaller coils that are positioned around the object being imaged. Compared to imaging with a single-channel volume coil, PA imaging allows for increased signal-to-noise ratio (SNR) and scan time reductions through parallel MRI (pMRI) techniques [1, 2]. Combining signal from individual coils such that their composite SNR is maximized requires knowledge of each coil’s complex-valued spatial sensitivity profile (also called sensitivity map or B1 map), which can easily be derived from the image of a reference volume coil (e.g., a body coil) with homogeneous sensitivity across the field-of-view (FOV) [1-3]. However, such reference coils are not always used alongside PA coils on high-field MRI scanners—especially on non-clinical systems—prompting the development of many algorithms that can estimate sensitivity information directly from the PA data itself. These algorithms vary dramatically in their assumptions about the complex-valued MR signal and, consequently, in the quality of their output images [4-8]. Thus, choosing the right coil combination algorithm is non-trivial for some applications [4, 6, 8].

Broadly, combination algorithms that estimate sensitivity information without using a physical volume coil fall into two categories: those that estimate each coil’s complex sensitivity map, and those that only estimate each coil’s phase offset (called “phase-matching” algorithms). Adaptive coil combination [9] approximates complex coil sensitivities through eigen-analysis of the coil array’s signal and noise covariance statistics within local regions of the image FOV. Sensitivities approximated in this manner are only defined up to multiplication with an arbitrary complex number—a scaling ambiguity that is typically resolved by referencing each coil’s estimated phase sensitivity to that of an arbitrary coil in the array. In regions where the reference coil has a low SNR, this approach creates image artifacts known as open-ended fringe lines in the coil-combined phase image [10-12].

The Virtual Reference Coil (VRC) algorithm [3] is computationally simple to implement and produces high-quality output [4, 6, 8, 13]. VRC requires the selection of a scalar value to which the phase of each coil is referenced, usually chosen as the mean phase within a region centered in the image or around a voxel in which coil sensitivities maximally overlap [4, 6, 14, 15]. More recently, “A Simple Phase Imaging Reconstruction method” (ASPIRE) [13] proposed to approximate each coil’s phase offset by exploiting the assumed linear evolution of phase between two echo times of a multi-echo Gradient Recalled Echo (GRE) sequence.

Prior work has also shown the utility of Singular Value Decomposition (SVD) based coil combination [11, 12, 16-20]. An SVD-based approach to sensitivity map estimation, called SVD-B1 (Fig 1) is evaluated here as an alternative to adaptive coil combination and phase matching algorithms conventionally used in QSM literature [5-8]. We assess SVD-B1’s effects on Quantitative Susceptibility Maps (QSM) of postmortem and in-vivo human brains. QSM is sensitive to the quality of its coil-combined magnitude and phase input [8, 21, 22] making it an ideal testbed for quantitatively evaluating the strengths and limitations of different coil combination algorithms. This study presents novel QSM images generated using SVD-B1 and employs a new “single-coil reference” technique to show that these images are generally more robust to noise and scan acceleration than those created using more conventional algorithms.

**FIG 1.**
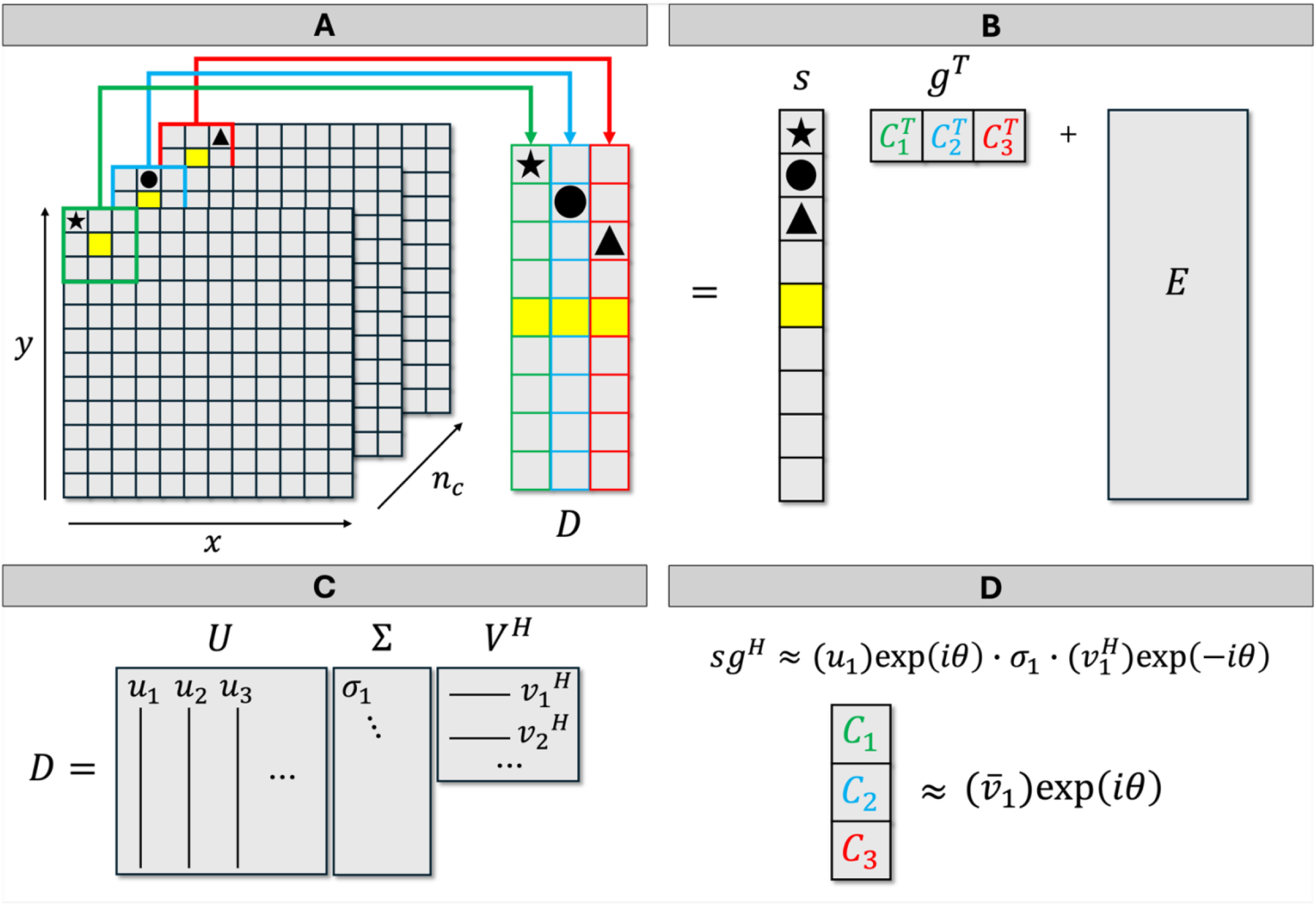
The Singular Value Decomposition B1 Algorithm: Schematic example of the Singular Value Decomposition (SVD)-B1 algorithm. (A) Image from *n*_*c*_ = 3 phased array coils, whose complex data are outlined in red, green, and blue respectively. The voxels within a 3 × 3 region of each coil’s complex image are rearranged to form the data matrix *D* as shown, with symbols denoting voxel positions. (B) *D* is assumed to be comprised of an additive noise component *E*, a signal component *s*, and a sensitivity component *g*, whose subcomponents (*C*_1_, *C*_2_, *C*_3_) correspond to the complex-valued sensitivity of each coil within the 3 × 3 image region. (C) The SVD of *D*. (D) The least-squares solution to the coil sensitivity vector is given 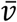, by the conjugate of *D*’s first right singular vector. The unit phase factor θ is a locally derived consensus phase across coils (the average phase of the first left singular vector *u*).

## Theory

Assume that the complex-valued sensitivity map of each PA coil *j* ∈ {1, …, *n*_*c*_} is constant over small *k*_*x*_ × *k*_*y*_ regions of a 2-dimensional image; this is a reasonable assumption in practice, since coil sensitivities are known to vary smoothly across the FOV (Erdogmus et al. 2004).

Given this assumption, the linear signal model for a single PA coil can be written as:

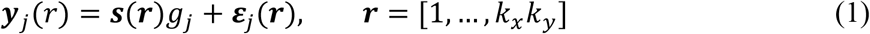

Where ***s***(***r***) is the image region’s MR signal vector, *g*_*j*_ is the coil’s complex-valued sensitivity, and *ε*_***j***_(***r***) is a vector of independent noise measurements. The complex-valued *k*_*x*_*k*_*y*_ × *n*_*c*_ MR data matrix *D* can be constructed as follows:

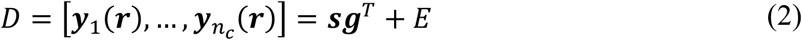

where *E* is a matrix of noise measurements which may be correlated across coils. Let *U*Σ*V*^*H*^ denote the SVD of *D*, with first left and right singular vectors ***u*** and ***v***, respectively. Given a pixel *m* centered in the *k*_*x*_ × *k*_*y*_ image region, the least-squares solution 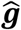 to *m*’s normalized coil sensitivity vector is given by the conjugate of ***v*** (Erdogmus et al. 2004; Sandgren et al. 2005; Bydder et al. 2008; Rodgers and Robson 2010; Inati, Hansen, and Kellman 2013, 2014).

However, ***v*** is only defined up to multiplication with an arbitrary complex number. This presents a scaling ambiguity in the estimated coil sensitivities, which can be resolved by referencing the phase of 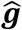 to a unit phase factor derived from ***u*** (Inati, Hansen, and Kellman 2013, 2014):

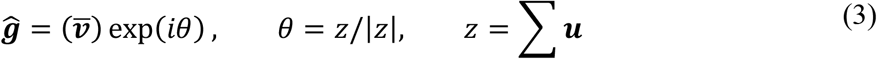

where 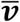 is the conjugate of ***v***. Repeating this calculation for each pixel in the image yields full sensitivity maps for all coils, which can then be used to combine the complex-valued coil images. This algorithm extends naturally to 3-dimensional image regions of size *k*_*x*_ × *k*_*y*_ × *k*_*z*_. A schematic example of the SVD-B1 algorithm is depicted in Figure 1.

## Materials and Methods

### Postmortem and In-Vivo Data

All imaging was conducted on a 7T MRI scanner (Siemens, Malvern Pennsylvania) equipped with a 32-channel head coil. MRI scans were performed on the standard spherical brain phantom (see Supplementary Fig 1), as well as a postmortem brain that was fixed in 10% neutral buffered formalin for two weeks prior to scanning. On the day of the MRI, the brain was transferred to a custom-built container filled with Fomblin as described in [23]. Imaging included a 3D multi-echo GRE sequence performed with: *TR* = 80ms; *TE* = 5ms, 10ms, 15ms, 20ms, 25ms, and 30ms; voxel size = 0.5mm × 0.5mm × 0.5mm; flip angle (FA) = 10°; image matrix size = 256 × 256 × 64. Coil-combined magnitude and phase images were acquired using both the sum of squares and adaptive coil combination algorithms available on the scanner. The raw coil array k-space data was saved for off-scanner reconstruction.

QSMs were also generated after adding Gaussian noise to each coil image in the fully sampled postmortem data. For each coil *c* and each echo time *j*, real and imaginary noise variances (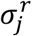 and 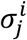) were approximated from noise-only image patches. Unique real and imaginary Gaussian noise profiles with variances 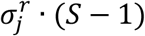 and 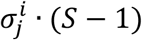 were added to the coil’s image (*c*_*j*_),approximately scaling its noise variances by the factor *S*. Five sets of coil volumes were created with progressively higher levels of noise, given by *S* = 5, 10, 20, and 40.

Additionally, QSMs were generated from images acquired with various levels of cartesian undersampling, given by the GRAPPA reduction factor *R* [24]. Postmortem images were acquired with *R* = 2, 3, 4, and 2 × 2, along with 24 reference lines for reconstruction; other imaging parameters were unchanged. Reconstructed images were acquired using the scanner’s implementation of GRAPPA and the adaptive combination algorithm. The raw coil array k-space data was saved for off-scanner reconstruction.

MRI was also performed on a healthy volunteer participant (male in their 70s), enrolled under the institutional review board-approved National Institute of Neurological Disorders and Stroke protocols “Evaluation of Progression in Multiple Sclerosis by Magnetic Resonance Imaging” (NCT00001248), after informed consent. A 3D multi-echo Gradient Recalled Echo (GRE) sequence was performed with the following parameters: *TR* = 50ms; *TE* = 5ms, 10ms, 15ms, 20ms, 25ms, and 30ms; voxel size = 1mm × 1mm × 1mm; *FA* = 10°; image matrix size = 168 × 192 × 64, with and without GRAPPA acceleration factor *R* = 2 × 2 and 24 reference lines. The “noisy” dataset for the in-vivo study was constructed with noise levels corresponding to *S* = 10, 20, 40, and 80. Additionally (see Supplementary Fig 2), high-resolution 2D GRE scans with TR = 1320 ms; TE=15, 32 ms; in-plane resolution of 0.1 mm; slice thickness = 1.1 mm and *R* = 2 were acquired on a participant (male in their 60s) of “thinking and memory problems in virologically controlled people living with HIV” protocol (NCT01875588) after informed consent.

### Off-Scanner Image Processing

All off-scanner reconstructions were implemented in MATLAB R2022a, Python 3, or Gadgetron [25]. The GRAPPA auto-calibration scan (ACS) and estimated sensitivity maps from the first *TE* were reused for reconstruction of all *TE*s to avoid using the noisy data at later echo times [15, 21].

#### Coil Combination

Coil volumes were combined into composite magnitude and phase images using the following coil combination algorithms:

##### SVD-B1 Combination

The SVD-B1 algorithm was implemented in Gadgetron using a kernel size of 7 × 7 × 5 or ∼2.5-3.5mm (SVD-B1). Since Gibbs ringing artifacts in individual coil images violate the assumptions of equation (1), sensitivity maps were estimated from data that was filtered by a symmetric Hann window function.

##### Combination Without Complex Sensitivity Maps

Coil magnitudes were combined offline in a root-sum-of-squares (RSS). Phases were combined offline in a magnitude-weighted sum after being phase matched by one of the following algorithms: **Hammond** [14], **VRC** [13], or **ASPIRE** [13] (implemented using the first two echoes).

Complex composite images were converted to the frequency domain and multiplied with a sinusoidal window function to emulate the on-scanner reconstruction pipeline’s smoothing function. Finally, a binary signal mask was created from each composite magnitude image by applying Gaussian smoothing (σ = 3) to the earliest echo time, followed by a high-pass binary filter with the threshold set to 20% of the maximum magnitude. Masks created from the baseline data (i.e., the fully sampled acquisition with no added noise) were reused in QSM processing across all levels of noise and acceleration.

#### Reconstruction of Accelerated Data

Undersampled k-space volumes were reconstructed using the following parallel MRI algorithms.

##### GRAPPA + ASPIRE, VRC, and Hammond

Individual coil images were reconstructed in Python using the PyGrappa library’s (v0.26.2, https://github.com/mckib2/pygrappa, [Accessed: November 11^th^, 2025]) Multidimensional GRAPPA (MDGRAPPA) function with parameters set as follows: kernel size = 5 × 4 × 4, regularization λ = 5 × 10^−4^. Sensitivity estimation was performed directly on the reconstructed coil volume (in accordance with [24]) using the ASPIRE, VRC, and Hammond algorithms for a more direct comparison between these three methods.

##### GRAPPA + SVD-B1

GRAPPA reconstruction weights were acquired using the GRAPPA functionality in Gadgetron’s Generic Reconstruction Chain with parameters set as follows: kernel size = 5 × 4 × 4, regularization threshold = 5 × 10^−4^, calibration overdetermination ratio = 45, and no coil compression. Since the presence of residual aliasing artifacts [26] in GRAPPA-reconstructed coil images violates the assumptions of equation (1), coil sensitivities were estimated from the ACS data (rather than directly from the coil images) using SVD-B1. To accomplish this, the ACS data was transformed to the image domain after being zero-padded to the full image matrix size and multiplied with a symmetric Hann window function to reduce Gibbs ringing artifacts. These sensitivities were used in tandem with Gadgetron’s GRAPPA reconstruction weights to simultaneously reconstruct and combine coil images in the manner described in [27] (SVD-B1 GRAPPA).

#### Reference Images and Regions-of-Interest

Partial ROIs containing the substantia nigra (SN), red nucleus (RN), left and right globus pallidus (GPL and GPR), and corticospinal tract (CSTL and CSTR) of the in-vivo and postmortem brains were manually delineated across multiple image slices. These ROIs were selected for their high-contrast susceptibility features associated with iron accumulation or dense myelination. ROI sizes ranged from 172-2744 voxels in the in-vivo brain and 1020-2585 voxels in the postmortem brain, with each ROI covering multiple, continuous slices. Within each ROI, a QSM was generated from the magnitude and phase image of the individual receiver coil with the highest SNR. Since this QSM was unbiased by the effects of any specific coil combination algorithm, it was used as a reference for comparison between QSMs generated using the different algorithms.

#### QSM Processing

All QSMs were generated using SEPIA’s [28] (version 1.2.2.6) “one-stop processing” pipeline, with: Echo Phase Combination = Optimum weights [4]; Phase Unwrapping = Laplacian (MEDI) [29, 30]; Background Field Removal (BFR) = VSHARP with 1 voxel of erosion before BFR, maximum radius = 10 voxels, and minimum radius = 3 voxels [30]. After BFR, a 4^th^ order 3D polynomial was removed from the tissue field as a precaution against residual B1 contributions [28, 31, 32]. Dipole inversion was performed using the MEDI algorithm with lambda = 1,000, edge mask threshold = 90, and no reference tissue [33-35].

### Statistical Analysis

Intraclass Correlation Coefficients (ICCs) were computed in 3D for each ROI to assess: (a) the consistency of each method across levels of data perturbation (perturbation agreement, ICC-PA), and (b) the agreement between the ROI’s single-coil reference QSM and QSMs generated by the various algorithms at each perturbation level (reference agreement, ICC-RA) [36]. Moreover, at each level of data perturbation, repeated-measures analysis of variance (RM-ANOVA) was performed to compare the mean susceptibilities of the various QSMs within each ROI. A heterogeneous compound symmetry (CSH) covariance structure, which allows for unequal variances across methods, was selected based on Akaike’s Information Criterion (AIC). Pairwise comparisons among methods were conducted using Tukey’s adjustment for multiple testing.

## Results

### SVD-B1 Yields the Most Noise-Robust QSMs

In-vivo phase images created using the MRI scanner’s adaptive combination algorithm contained open-ended fringe lines within central slices of the image; these artifacts corresponded to regions of lost signal in the magnitude (wormhole artifacts, Fig 2A), usually contained large background field effects, and produced erroneous QSM features. Compared to phase images produced by other off-scanner algorithms, those produced by SVD-B1 displayed reduced background field effects and fewer phase wraps. No clearly erroneous features were visible in magnitude or QSM images produced with any of the off-scanner algorithms. In-vivo QSMs generated from individual coils with the highest SNR in the region containing the scanner-generated artifacts (coils #11 and #15; Fig 2F-G) did not contain the erroneous features, corroborating the off-scanner reconstructions. Since coil #15 (Fig 2G) had the highest SNR within the image region containing the scanner-generated artifact (Fig 2, red box), its QSM was chosen as the region’s reference dataset. ICC-RA within the artifact region was highest for coil #11’s QSM and lowest for the QSM produced from scanner-generated images (Fig 2H), with no 95% CI overlap between these two ICC values. In the postmortem data (not shown), scanner-generated magnitude, phase, and QSM images were likewise corrupted by artifacts absent from single-coil images and images created using off-scanner algorithms.

**FIG 2.**
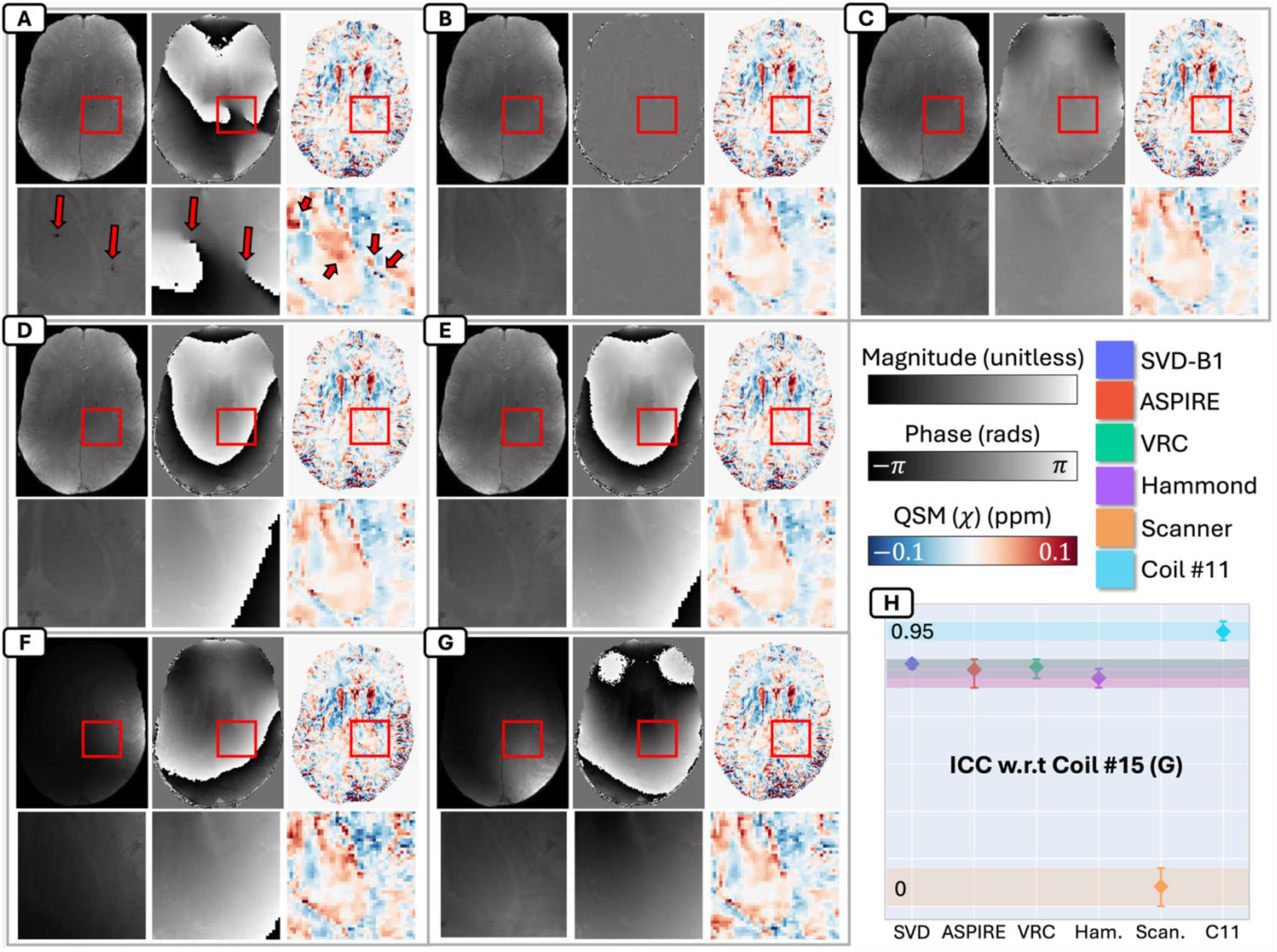
Qualitative differences in in-vivo images and Quantitative Susceptibility Maps (QSM) reconstructed from baseline data using various coil combination algorithms: Representative slice of a 3D GRE in-vivo acquisition showing, from left to right in each subpanel, magnitude, phase, and QSM from whole slice (top row) and a zoomed-in region (red box, bottom row) generated using (A) the adaptive combination algorithm available on the scanner; (B) the Singular Value Decomposition (SVD)-B1 algorithm; (C) ASPIRE, (D) VRC, and (E) the Hammond algorithm. Red arrows indicate artifacts in the magnitude and phase images. (F) Images generated from coil #11, which had the 2^nd^ highest signal-to-noise ratio of all coils within the zoomed region. (G) Images generated from coil #15, which had the highest signal-to-noise-ratio of all coils within the zoomed region. (H) Intraclass Correlation Coefficients (ICC) measuring agreement between each QSM and the QSM of coil #15. Error bars and shaded regions denote ICC 95% confidence intervals.

The quality of postmortem QSMs decreased as progressive levels of Gaussian noise were added to the uncombined coil data. At the maximum level of added noise (*S* = 40), postmortem magnitude and QSM images created using SVD-B1 (Fig 3A, top row) were visibly less noisy, especially near the edges of the tissue, than those created using other algorithms (Fig 3A, second to fourth rows). Magnitude images created using the Hammond algorithm contained notable speckling artifacts at *S* = 40, which caused severe QSM errors near the edges of the tissue (Fig 3A, bottom row). Of all postmortem QSMs, those created using SVD-B1 achieved the highest ICC-PA across noise levels (Fig 3B) by 1.1–13.3% across the SN, RN, GPL, GPR, and CSTL, with no overlap in 95% CIs. In the CSTR, ICC-PA was similar (i.e., 95% CIs almost completely overlapped) for SVD-B1 (95% CI=0.92–0.94), ASPIRE (0.937–0.94), and VRC (0.937–0.94), respectively. Of all postmortem QSMs produced from *S* = 40 data, those created using SVD-B1 achieved the highest ICC-RA (Fig 3C) by 36.6% in the GPL, with no overlap in corresponding CIs. In all other ROIs, these 95% CIs overlapped. At *S* = 40, QSMs generated using SVD-B1 significantly differed from other algorithms’ QSMs (i.e., *p* < 0.05 for the difference in mean susceptibilities, as measured by RM-ANOVA) within all ROIs except the RN (where *p* > 0.05 for SVD-B1/ASPIRE and SVD-B1/VRC).

**FIG 3.**
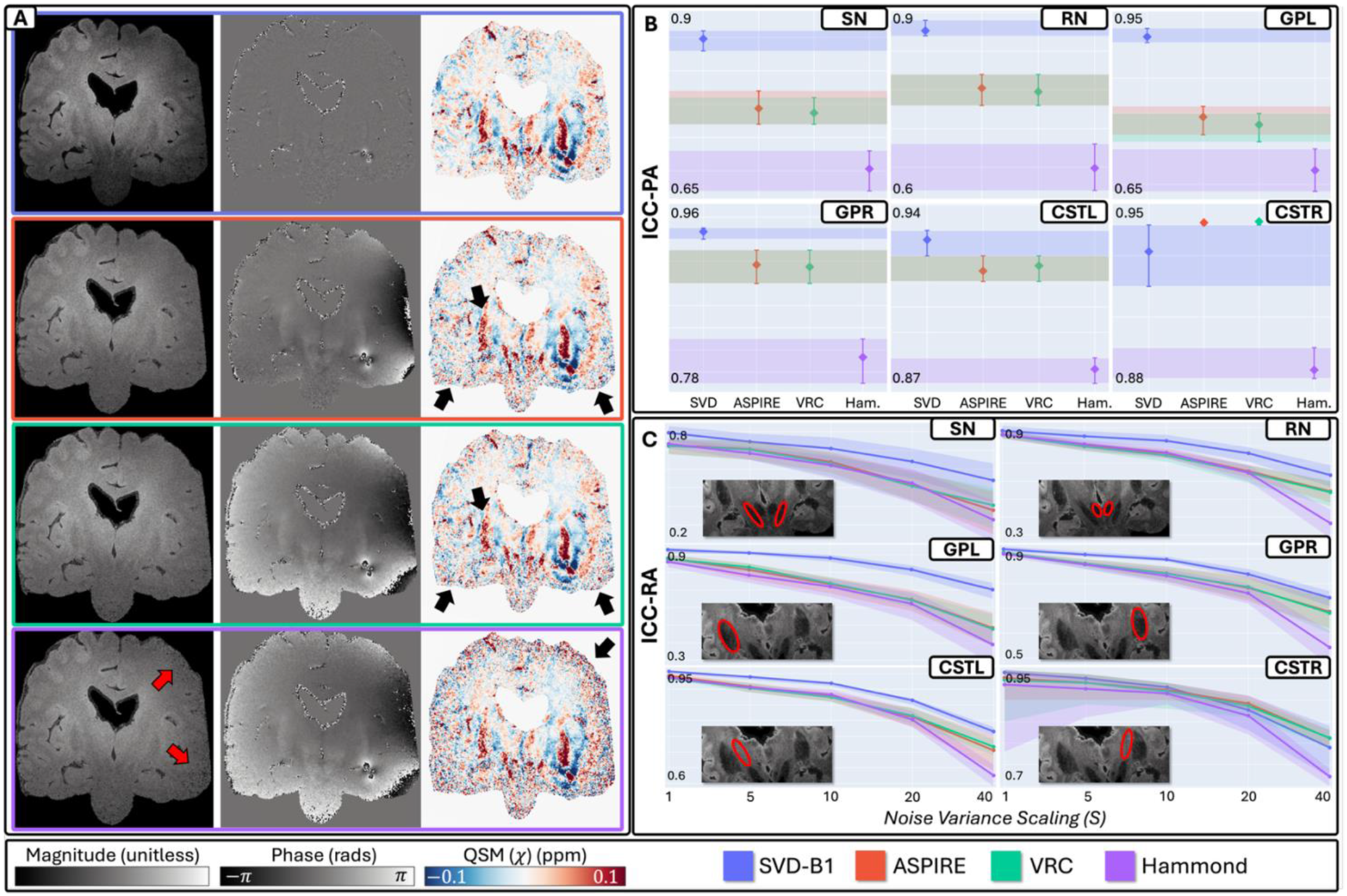
Effect of noise on postmortem images and Quantitative Susceptibility Maps (QSM) reconstructed using various coil combination algorithms: (A) A representative slice from a 3D GRE acquisition of a postmortem brain. From left to right in each subpanel: magnitude, phase, and QSM images created from coil data with the maximum level of added noise (*S* = 40) using the Singular Value Decomposition (SVD)-B1 algorithm (top row), ASPIRE (second row), VRC (third row), and Hammond algorithm (bottom row). Black arrows indicate regions where noise levels visibly differ between the QSMs in each subpanel and red arrows indicate magnitude image artifacts. (B) The Intraclass Correlation Coefficients measuring QSM agreement across noise levels (ICC-PA). (C) Each QSM’s Intraclass Correlation Coefficient with the single-coil reference data (ICC-RA) selected for the following regions-of-interest (ROIs): Substantia Nigra (SN), Red Nucleus (RN), Globus Pallidus Left (GPL), Globus Pallidus Right (GPR), Corticospinal Tract Left (CSTL), and Corticospinal Tract Right (CSTR). *S* = 1 corresponds to the baseline data (i.e., the fully sampled coil volume with no added noise). Error bars and shaded regions denote ICC 95% confidence intervals.

The in-vivo data yielded similar results to the postmortem data. At *S* = 80, in-vivo magnitude and QSM images created using SVD-B1 (Fig 4A, top row) were visibly less noisy near the edges of the tissue than those created using other algorithms (Fig 4A, second to fourth rows). As in the postmortem data, images created using the Hammond algorithm (Fig 4A, bottom row) contained notable speckling artifacts that propagated errors to QSM. Of all in-vivo QSMs, those created using SVD-B1 achieved the highest ICC-PA (Fig 4B) by 1.76–3.1% across all ROIs, with no 95% CI overlap. Moreover, of all QSMs produced from *S* = 80 data, those created using SVD-B1 achieved the highest ICC-RA by 8.5% in the GPL and 3.5% in the GPR, with no overlap in corresponding 95% CIs. In all other ROIs, these 95% CIs overlapped, though SVD-B1’s QSMs consistently achieved the best ICC-RA lower-bound. At S=80, SVD-B1’s QSM significantly differed (*p* < 0.05) from other algorithms’ QSMs within the CSTL, CSTR, and GPL. In the SN, RN, and GPR, SVD-B1’s QSM was similar (i.e., *p* > 0.05) to those generated by other algorithms (*p* > 0.05 for SVD-B1/ASPIRE and SVD-B1/VRC in the GPR; SVD-B1/ASPIRE, SVD-B1/Hammond in the RN; SVD-B1/VRC in the SN).

**FIG 4.**
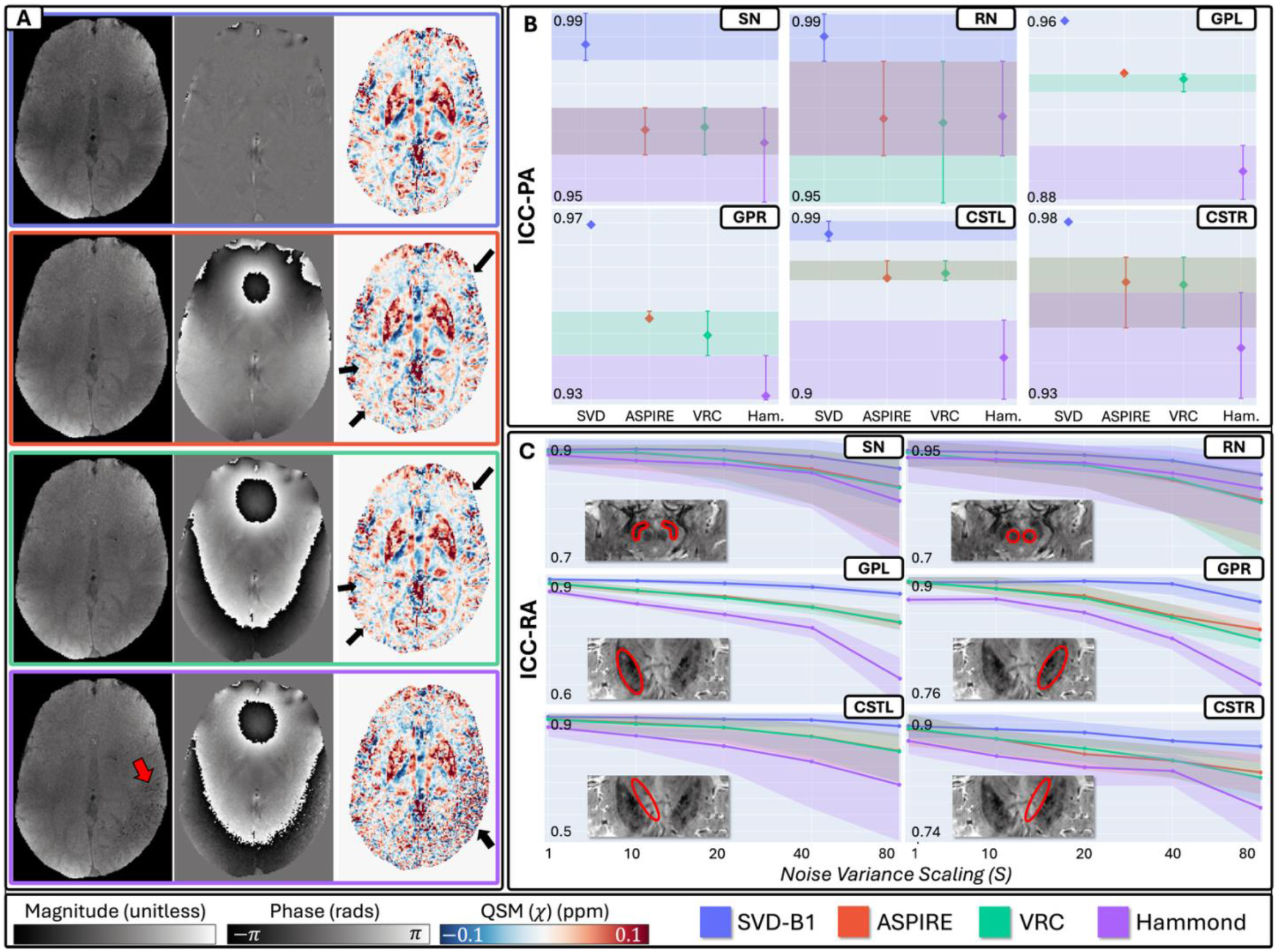
Effects of noise on in-vivo images and Quantitative Susceptibility Maps (QSM) reconstructed using various coil combination algorithms: (A) Representative slice from a 3D GRE acquisition of an in-vivo brain. From left to right in each subpanel: magnitude, phase, and QSM images created from coil data with the maximum level of added noise (*S* = 80) using the Singular Value Decomposition (SVD)-B1 algorithm (top row), ASPIRE (second row), VRC (third row), and Hammond algorithm (bottom row). Black arrows indicate regions where noise levels visibly differ between the QSMs in each subpanel, and red arrows indicate magnitude image artifacts. (B) The Intraclass Correlation Coefficients measuring QSM agreement across noise levels (ICC). (C) Each QSM’s Intraclass Correlation Coefficient with the single-coil reference data (ICC-RA) selected for the following regions-of-interest (ROIs): Substantia Nigra (SN), Red Nucleus (RN), Globus Pallidus Left (GPL), Globus Pallidus Right (GPR), Corticospinal Tract Left (CSTL), and Corticospinal Tract Right (CSTR). *S* = 1 corresponds to the baseline data (i.e., the fully sampled coil volume with no added noise). Error bars and shaded regions denote ICC 95% confidence intervals.

### SVD-B1 GRAPPA Yields Acceleration-Robust QSMs

Qualitative analysis revealed that QSMs generated from rapidly acquired data became noisier as scan acceleration (*R*) increased (Fig 5). Phase images produced from the *R* = 2 × 2 postmortem data using the MRI scanner’s reconstruction pipeline contained open-ended fringe lines that propagated severe errors to QSM (Fig 5A, third row from top). While these artifacts were present in all scanner-generated phase images (data not shown), their effects seemed to compound with the effects of other artifacts and of noise amplification at *R* = 2 × 2, resulting in a particularly low quality QSM. No such errors were present in postmortem images created using the off-scanner algorithms. At *R* = 2 × 2, postmortem QSMs were visibly less noisy when created with SVD-B1 GRAPPA (Fig 5B, third row from top) than with all other algorithms (Fig 5A, C-E, third row from top). At *R* = 4, QSMs were least noisy when created with SVD-B1 GRAPPA and, surprisingly, the on-scanner algorithm (Fig 5, bottom row). The fact that—in contrast to other algorithms—the on-scanner algorithm performed better at *R* = 4 than *R* = 2 × 2 likely owes to the specific coil geometries within the scanner and to additional, unknown processing steps in the scanner’s reconstruction pipeline.

**FIG 5.**
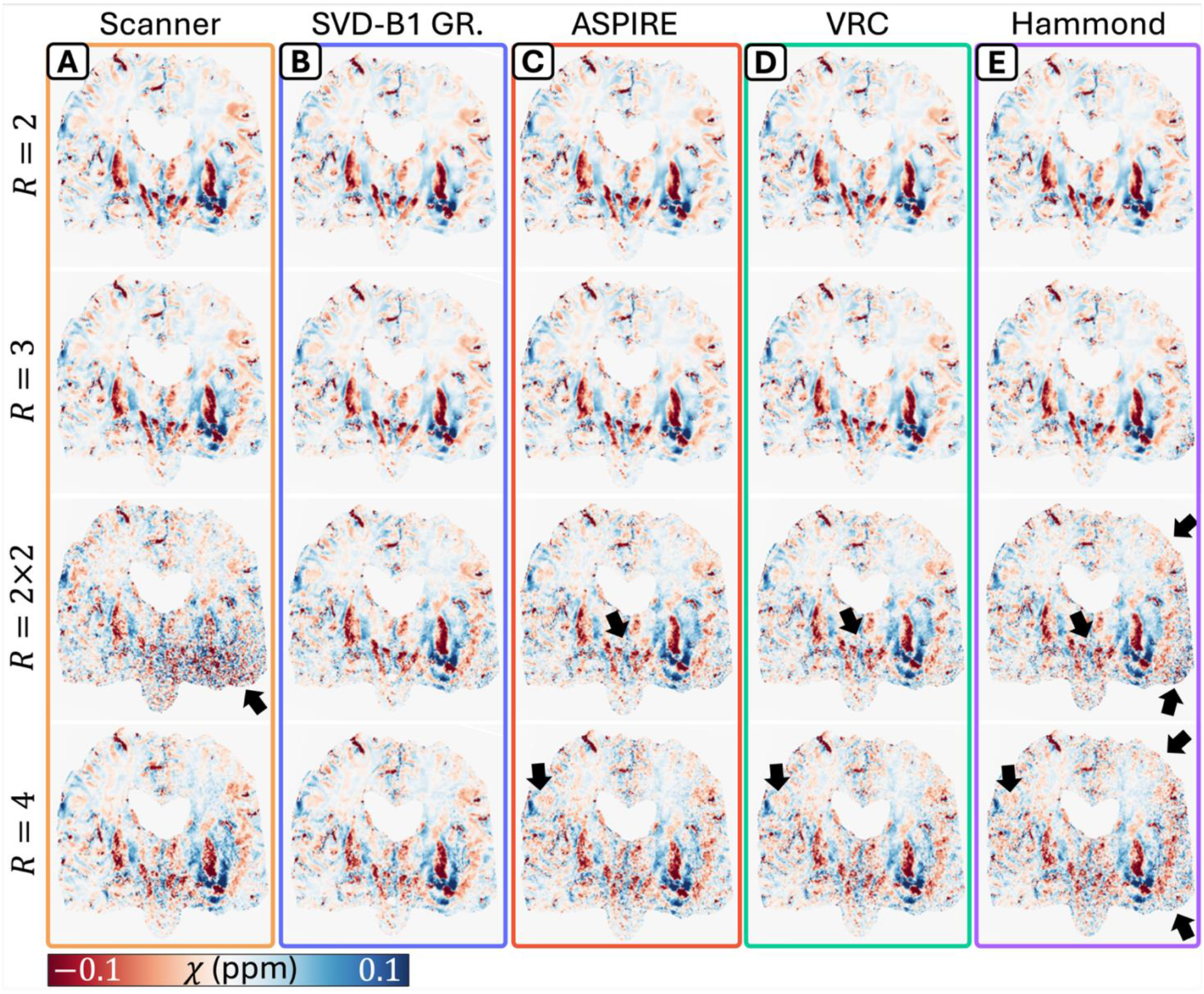
Qualitative effects of acceleration on postmortem Quantitative Susceptibility Maps (QSM): QSM images generated from undersampled data—GRAPPA acceleration factor *R* = 2, 3, 2 × 2, and 4—generated using: (A) the on-scanner GRAPPA algorithm, (B) SVD-B1 GRAPPA, (C) GRAPPA + ASPIRE, (D) GRAPPA + VRC, and (E) GRAPPA + Hammond.

In-vivo phase images created using the scanner’s reconstruction pipeline likewise suffered from open-ended fringe lines at *R* = 2 × 2, which caused severe QSM errors throughout the tissue (Fig 6A). Such artifacts were mostly absent in off-scanner reconstruction algorithms, except in regions very close to large field perturbations such as just above the frontal sinuses. In contrast to the postmortem results, no visible difference in quality was observed between in-vivo QSMs generated by the various off-scanner algorithms at any acceleration level. Correspondingly, 95% CIs for the ICC-PA and -RA metrics almost completely overlapped in all in-vivo ROIs (data not shown). At *R* = 2 × 2, in-vivo QSMs produced using SVD-B1 GRAPPA, ASPIRE, VRC, and Hammond were similar (*p* > 0.05) within all ROIs except the CSTR (where *p* < 0.05 for SVD-B1 GRAPPA/ASPIRE, SVD-B1 GRAPPA/VRC, and SVD-B1 GRAPPA/Hammond). Note that the reference coil SNRs in the fully sampled in-vivo scan were approximately twice those of the postmortem scan.

**FIG 6.**
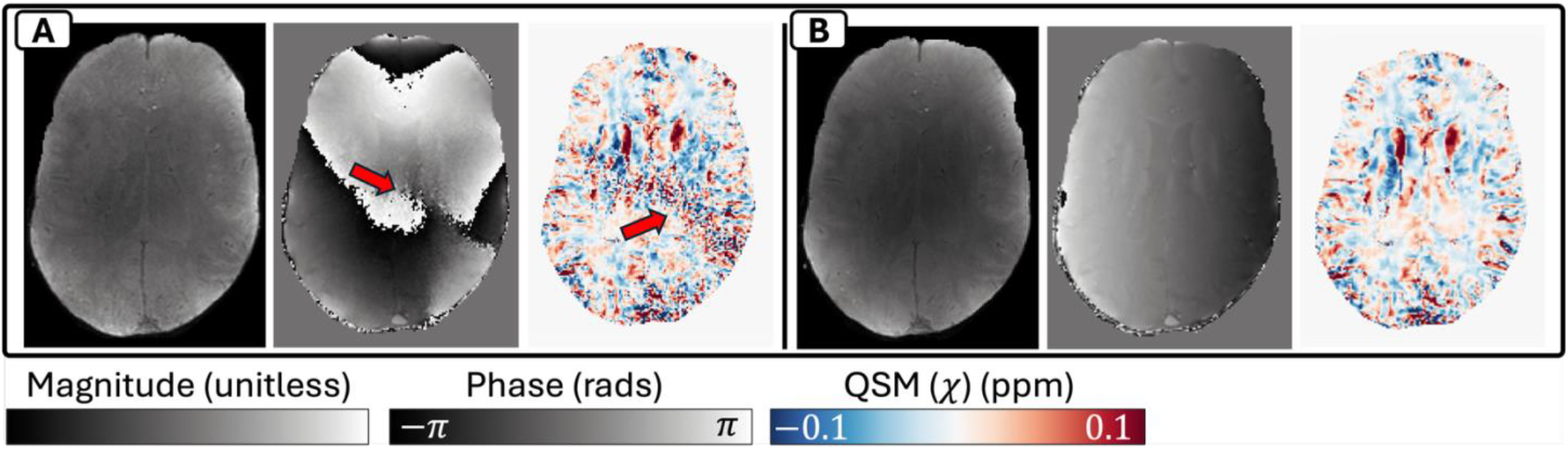
Effects of acceleration on in-vivo Quantitative Susceptibility Maps (QSM): From left to right in each subpanel, magnitude, phase, and QSM images are shown, reconstructed from *R* = 2 × 2 undersampled data using (A) the on-scanner GRAPPA algorithm and (B) the SVD-B1 GRAPPA algorithm. Red arrows indicate image artifacts.

Of all postmortem QSMs, those created using SVD-B1 GRAPPA achieved the highest ICC-PA across acceleration levels (Fig 7A) by 7.9–31.2% across the SN, RN, GPR, CSTL, and CSTR, with no overlap in 95% CIs. In the GPL, ICC-PA values were similar for SVD-B1 GRAPPA (95% CI = 0.72–0.77), ASPIRE (0.67–0.73), and VRC (0.65–0.72). Postmortem QSMs created using SVD-B1 GRAPPA also exhibited the highest ICC-RA at *R* = 2 × 2 by 20.1% in the CSTL, with no 95% CI overlap. Likewise, at *R* = 4, these QSMs achieved the highest ICC-RA by 4.8% in the CSTL and 23.4% in the GPR. For all other ROIs and acceleration levels, 95% CIs overlapped between methods. At *R* = 2 × 2, SVD-B1 GRAPPA’s postmortem QSM significantly differed from other algorithms’ QSMs in all ROIs except the CSTL (where *p* > 0.05 for SVD-B1 GRAPPA/ASPIRE and SVD-B1 GRAPPA/Hammond). At *R* = 4, SVD-B1 GRAPPA’s QSM and the QSM generated using the on-scanner algorithm were similar (*p* > 0.05) within the SN, RN, and GPR.

**FIG 7.**
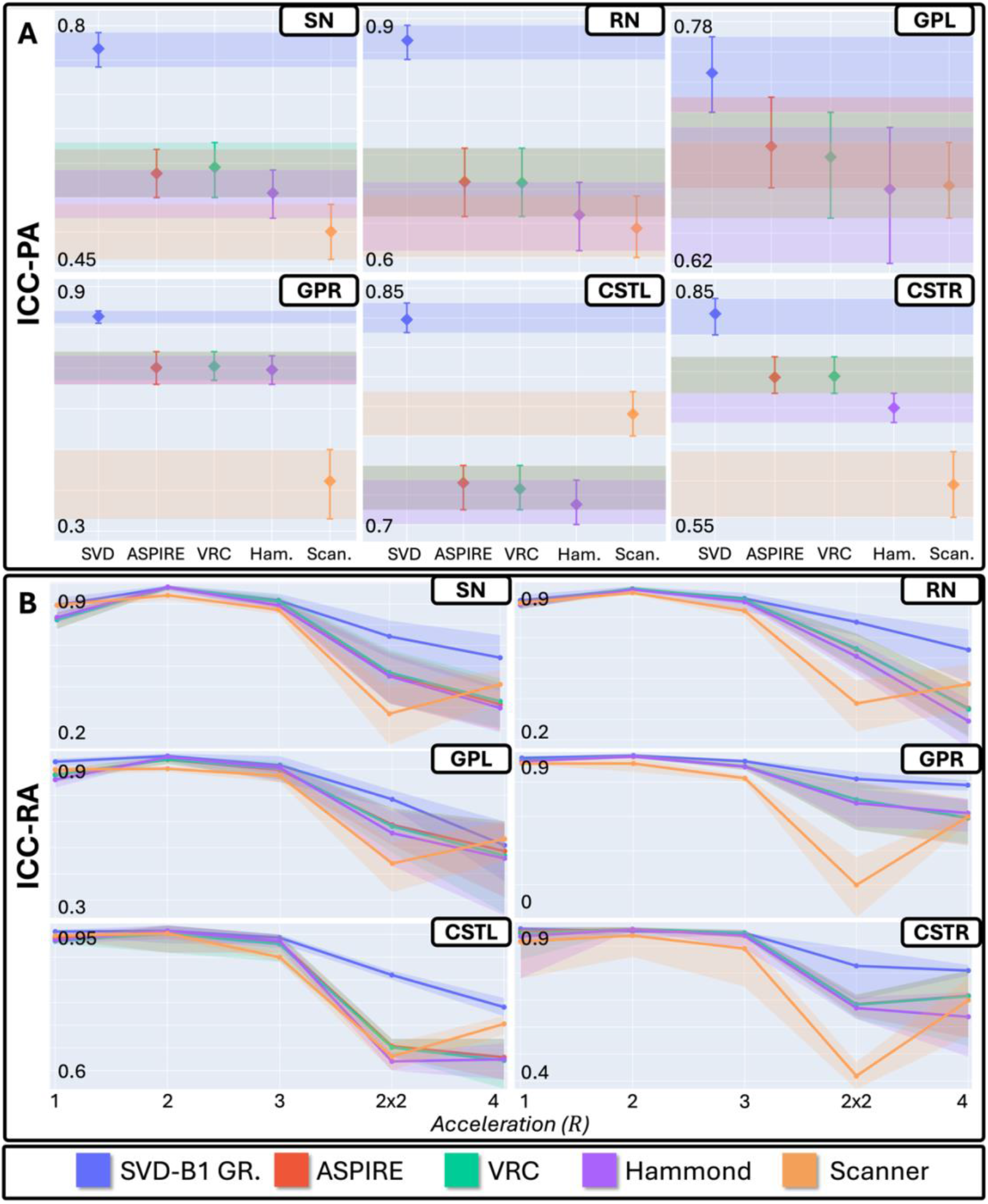
Quantitative effects of acceleration on postmortem Quantitative Susceptibility Maps (QSM): (A) The Intraclass Correlation Coefficients measuring QSM agreement across acceleration levels (ICC), with error bars and shaded regions denoting ICC 95% confidence intervals. (B) Each QSM’s Intraclass Correlation Coefficient with the single-coil reference data (ICC-RA) for the following regions-of-interest: the Substantia Nigra (SN), Red Nucleus (RN), Globus Pallidus Left (GPL), Globus Pallidus Right (GPR), Corticospinal Tract Left (CSTL), and Corticospinal Tract Right (CSTR). *R* = 1 corresponds to the baseline data (i.e., the fully sampled coil volume). Error bars and shaded regions denote ICC 95% confidence intervals.

Supplementary Figure 1 shows contiguous slices of a standard Siemens spherical phantom in which the behavior of the scanner-generated phase artifact is clearly depicted in a homogenous sample. Supplementary Figure 2 depicts a clinical case where the on-scanner reconstruction produced a wormhole artifact that could be clinically misinterpreted, whereas offline SVD-B1 reconstruction did not.

## Discussion

Algorithms that combine complex coil images without separately acquired coil sensitivity maps vary in the quality of their output magnitude and phase images, which can, in turn, affect the accuracy and noise-robustness of QSM. Of the reconstruction pipelines compared in this study, those using SVD-B1 and its parallel imaging extension, SVD-B1 GRAPPA, generally produced the most robust and accurate QSMs from noisy and rapidly acquired data. Similar to adaptive coil combination [9], SVD-B1 estimates complex coil sensitivities from the PA data itself. However, unlike adaptive combination, SVD-B1 accomplishes this without referencing each coil’s phase sensitivity to that of another, potentially low SNR coil in the array. Unlike other conventional algorithms, SVD-B1 neither relies on a global scalar reference phase nor assumes that phase evolves linearly over time. Unlike ASPIRE, it does not rely on multi-echo data, so can potentially enable faster acquisitions through *TR* reduction. By referencing each coil’s phase sensitivity to a locally derived consensus phase across coils (the average phase of the left singular vector within each image region), SVD-B1 produces phase images with relatively few wraps and reduced background field effects. Our findings suggest SVD-B1 and SVD-B1 GRAPPA as algorithms of choice for high-resolution imaging at clinically acceptable scan times.

In this study, noise dominated large regions of in-vivo and postmortem images created using the Hammond algorithm. These artifacts likely owed to the spatial variance of phase offset across the FOV, which is known to cause phase matching errors in image regions far from the selected phase reference ROI [4, 15]. Magnitude and phase images created using ASPIRE and VRC contained no such artifacts, but their resulting QSMs were generally inferior to those created using SVD-B1 whenever high levels of noise were present in the uncombined coil data.

Accurate image reconstruction is desirable for multiple reasons. QSM has recently shown promise in the study of neurological disorders such as Parkinson’s disease [37-39] and multiple sclerosis [40-43], and can help quantify microstructural changes. Phase images are increasingly being used to detect subtle pathological features in the brain, such as paramagnetic rim lesions in multiple sclerosis [44, 45]. Even magnitude images are affected by coil combination inaccuracies—e.g., magnitude wormhole artifacts can mimic or obscure clinically relevant tissue features such as microbleeds, venous malformations, or ventral veins in lesions. In this study, magnitude, phase, and QSM images produced using SVD-B1 were accurate and artifact free in the brain parenchyma when reconstructed under reasonable noise conditions and acceleration.

Our study did not use a phantom of known susceptibility to evaluate the accuracy of QSMs generated from coil-combined data [46]. Instead, accuracy was evaluated with respect to the QSM of a single “reference” coil, chosen for each ROI as the coil with the highest SNR. This QSM provided ROI-specific reference measurements that were unbiased by the assumptions of a digital phantom [8] or the effects of any specific coil combination algorithm [6]. For highly noisy and rapidly acquired data, the quantitative metrics acquired with this technique agreed with our qualitative results: e.g., large quantitative differences between the Hammond algorithm’s QSM and the reference data corresponded with the visibly low quality of Hammond’s QSMs. However, the “single-coil reference” technique is limited by the fact that QSMs generated from an individual coil are necessarily noisier than those created from optimally combined composite images [1]. This SNR difference introduces a slight bias into our study’s evaluation strategy.

This study did *not* aim to evaluate different approaches to QSM processing steps beyond coil combination, such as BFR and dipole inversion. All QSMs were generated using a generic SEPIA configuration. Other configurations and algorithms are possible—perhaps even preferable, depending on the context—but are beyond the scope of this work. For a comprehensive review of QSM processing methods, see [47]; for examples of other SEPIA configurations, see [28]. It is important to note that, while SVD-B1 GRAPPA was implemented using GRAPPA reconstruction weights computed in Gadgetron, other off-scanner algorithms used weights computed in PyGrappa due to technical limitations. Discrepancies between these two GRAPPA implementations (e.g., kernel formation techniques, regularization methods, etc.) likely also contributed to the differences observed between QSMs created from accelerated data, but this was not explored herein. Nevertheless, QSMs generated from fully sampled data (i.e., when no GRAPPA reconstruction was required) using SVD-B1 were of a notably higher quality than those produced using other algorithms.

Where possible, reconstruction algorithms were configured per their official implementations and/or their implementations in prior literature. For example, the phase offset computed by ASPIRE was smoothed using the default “approximate Gaussian filter” implemented in the algorithm’s official repository [13]. To smooth the phase offset maps computed by VRC, we used a median filter in the manner of [6]. For concordance with [15], we used a 3 × 3 × 3 reference region for the Hammond and VRC algorithms. We experimented with various SVD-B1 kernel sizes ranging from 3×3×3 to 11×11×11, ultimately choosing a 7×7×5 kernel heuristically as a middle ground for sensitivity map smoothing. In contrast to SVD-B1 GRAPPA, in which sensitivity maps were estimated from the ACS data, the ASPIRE, Hammond, and VRC algorithms were applied directly to GRAPPA-reconstructed coil images in concordance with [24]. Changing these configurations (e.g., using different smoothing functions or kernel sizes) affected the resulting magnitude, phase, and QSM images. Finding the optimal configuration for each algorithm is grounds for future work but is beyond the scope of the present study.

In conclusion, we demonstrated that magnitude, phase, and QSM images produced using SVD-B1 based algorithms are generally more robust to noise and scan acceleration than those created using more conventional reconstruction algorithms. Applications using these techniques stand to benefit from accurate, noise-robust pMRI and high-resolution imaging to reveal fine-grained tissue structure that can be acquired at a clinically acceptable scan time.

## Data Availability

The data and code will be made available upon request, subject to NIH’s data sharing policies.

## Acknowledgements

This research was supported in part by the Intramural Research Program of the National Institutes of Health (NIH). The contributions of the NIH author(s) were made as part of their official duties as NIH federal employees, are in compliance with agency policy requirements, and are considered Works of the United States Government. However, the findings and conclusions presented in this paper are those of the author(s) and do not necessarily reflect the views of the NIH or the U.S. Department of Health and Human Services. We are deeply grateful to our patients and their families for their willingness to participate in organ donation and to authorize autopsy procedures for biomedical research. In addition, we would like to thank the Clinic staff who cared for the patients and facilitated their autopsies. Further, we would like to thank Jacco De Zwart for providing his expertise and the NIH fMRI core facility for imaging

