## Supplementary Figures for "Singular Value Decomposition-Based Coil Combination Improves the Accuracy and Noise-Robustness of Quantitative Susceptibility Maps"

### Supplementary Material

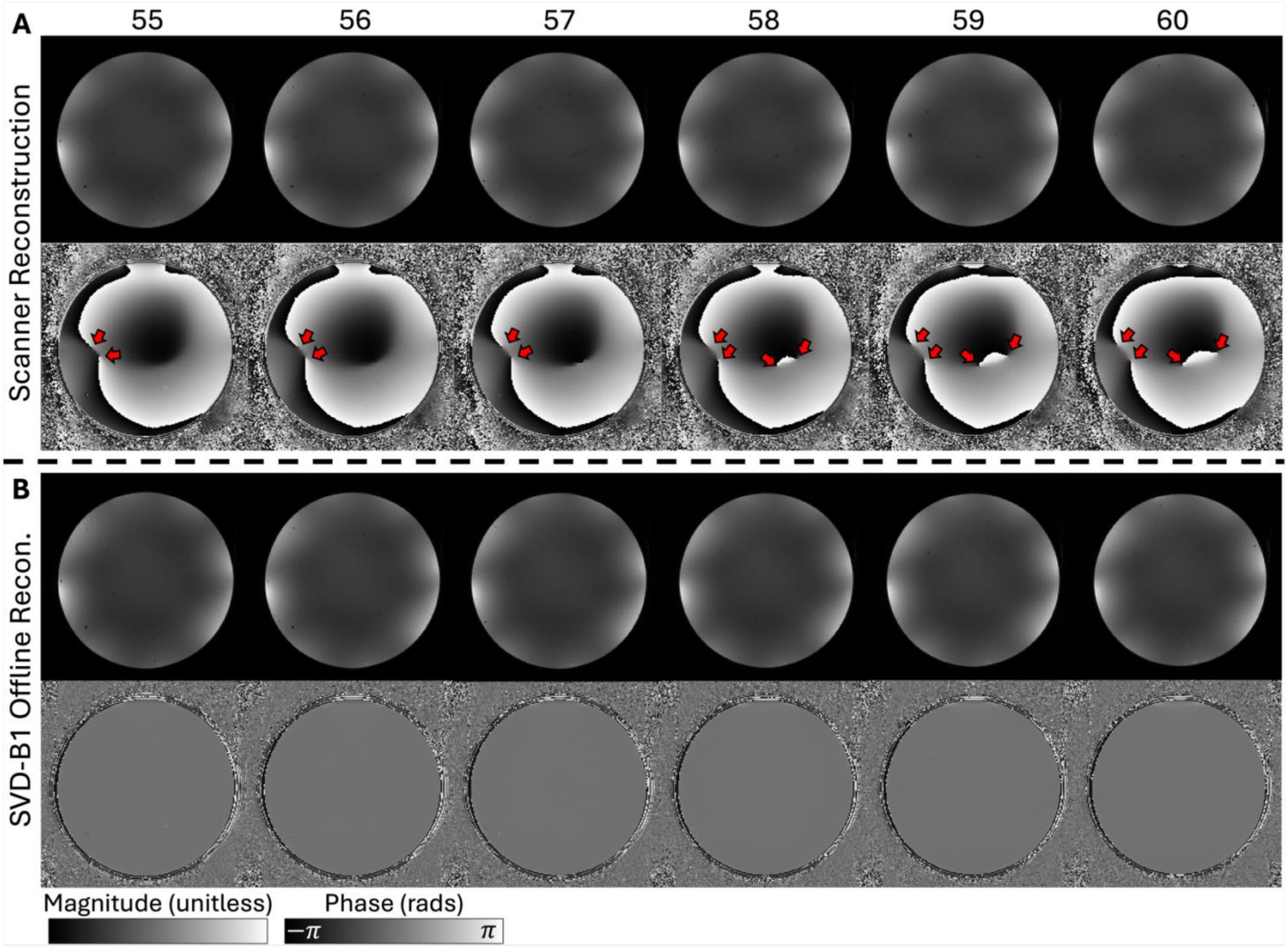


**SUPPLEMENTARY FIG 1:** Six contiguous slices (slices 55-60) from a standard Siemens spherical phantom acquired using a 3D GRE sequence. (A) Images reconstructed on the scanner. (B) Images reconstructed offline using the SVD-B1 algorithm. Open contour phase artifacts are clearly seen in phase images reconstructed by the scanner (arrows) but are absent in those reconstructed using SVD-B1. Magnitude images from the scanner also contain wormhole artifacts, which are too small to appreciate at this scale.

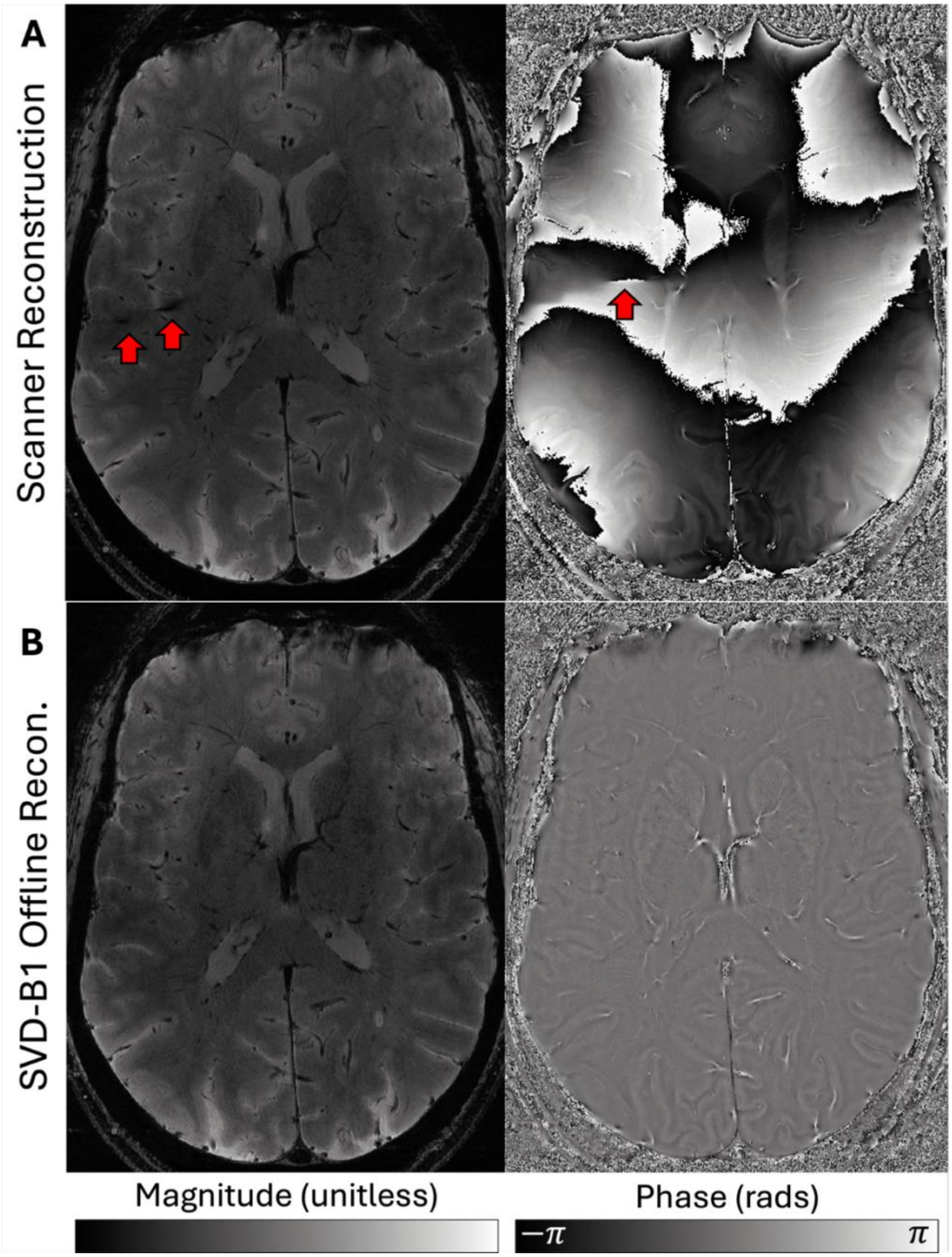


**SUPPLEMENTARY FIG 2:** High-resolution 2D GRE images of an in-vivo human brain (A) reconstructed on the scanner and (B) reconstructed using the SVD-B1 algorithm. The scanner-generated images contain open contour phase artifacts and magnitude wormhole artifacts (red arrows) that can be misinterpreted as blood vessels or microbleeds. Images reconstructed using SVD-B1 do not contain these artifacts. To create the SVD-B1 images, we experimented with 2D kernel sizes ranging from 7 x 7 to 40 x 40, ultimately selecting a 20 x 20 kernel heuristically, as a middle ground for sensitivity map smoothing. Note that the images above were much higher-resolution than individual slices of the in-vivo 3D GRE scan used in the main text (0.1mm inplane here vs. 0.5mm for in-vivo 3D GRE).
